## Supplementary figures and images for "Development and validation of the Japanese version of the Lesbian, Gay, Bisexual, and Transgender Development of Clinical Skills Scale"

### S1 Fig

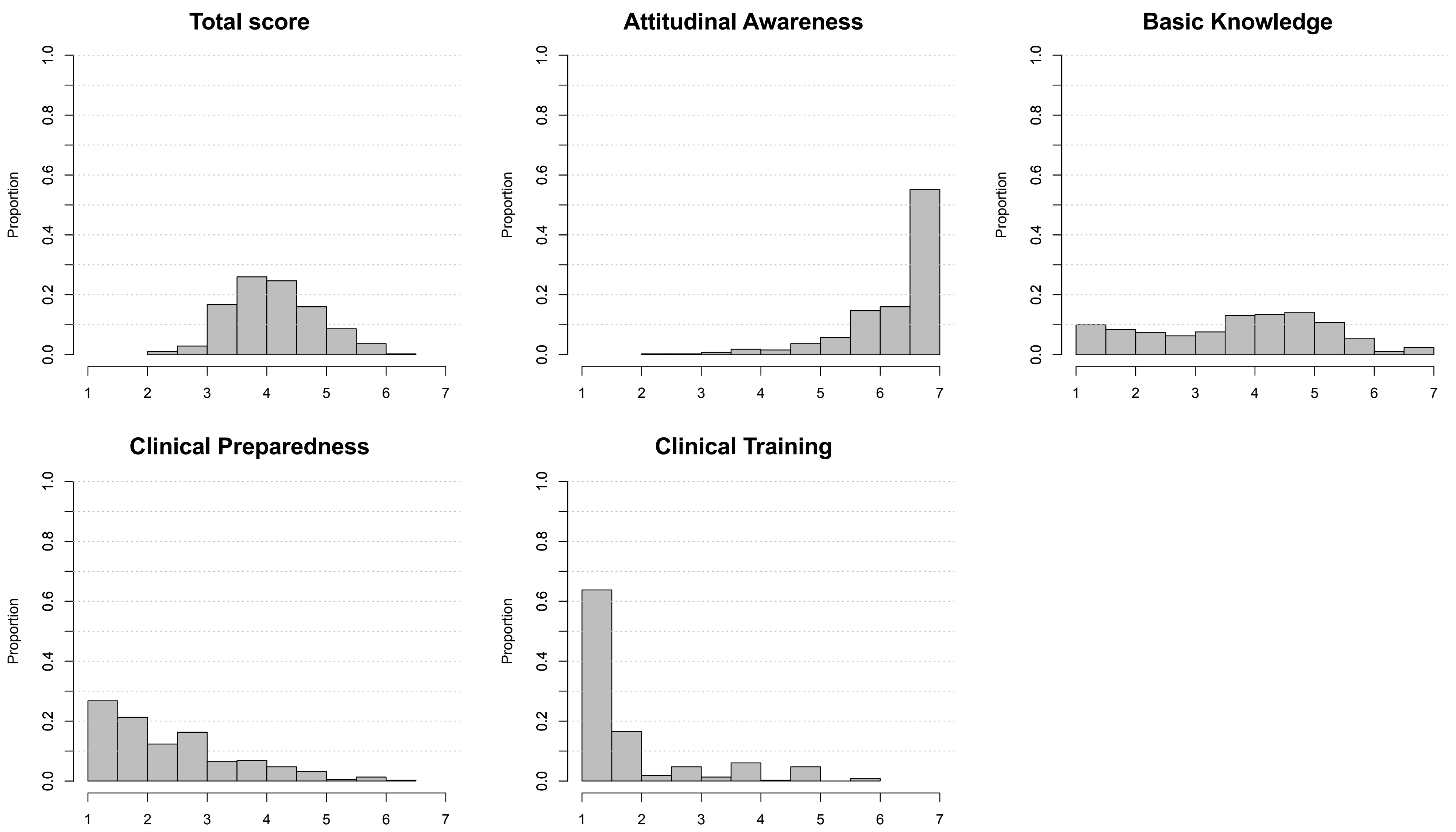


**S1 Fig. The score distributions for the overall and each subscale**
