## Supplementary material for "Development and validation of the Japanese version of the Lesbian, Gay, Bisexual, and Transgender Development of Clinical Skills Scale": S1 Table

**S1 Table. Hypothesis testing among cisgender-heterosexual participants (n = 293)**

| | No. | Total (mean (SD)) | Attitudinal Awareness (mean (SD)) | Basic Knowledge (mean (SD)) | Clinical Preparedness (mean (SD)) | Clinical Training (mean (SD)) |
|---|---|---|---|---|---|---|
| Homosexual friends/family | | | | | | |
| Yes | 41 | 4.31 (0.81) | 6.45 (0.97) | 3.78 (1.56) | 2.71 (1.38) | 1.93 (1.45) |
| Probably | 14 | 4.33 (0.57) | 6.44 (0.42) | 4.71 (1.29) | 2.19 (1.02) | 1.57 (1.00) |
| Probably not | 135 | 4.10 (0.72) | 6.28 (0.81) | 3.66 (1.51) | 2.33 (1.15) | 1.81 (1.30) |
| No | 103 | 3.95 (0.62) | 6.15 (0.89) | 3.59 (1.28) | 2.11 (0.99) | 1.55 (1.04) |
| P value | | **< 0.01** | **< 0.01** | 0.09 | **0.04** | 0.11 |
| Homosexual coworker | | | | | | |
| Yes | 74 | 4.45 (0.78) | 6.43 (0.8) | 4.07 (1.57) | 2.89 (1.31) | 2.23 (1.60) |
| Probably | 72 | 4.11 (0.69) | 6.28 (0.91) | 3.73 (1.39) | 2.38 (1.10) | 1.58 (1.08) |
| Probably not | 120 | 3.87 (0.61) | 6.13 (0.87) | 3.48 (1.40) | 1.96 (0.89) | 1.53 (0.99) |
| No | 27 | 4.00 (0.48) | 6.36 (0.71) | 3.62 (1.28) | 1.96 (1.06) | 1.59 (1.01) |
| P value | | **< 0.01** | **0.02** | **< 0.01** | **< 0.01** | **< 0.01** |
| Trans friends/family | | | | | | |
| Yes | 17 | 4.74 (0.89) | 6.46 (0.87) | 4.65 (1.27) | 3.08 (1.51) | 3.00 (1.87) |
| Probably | 11 | 4.44 (0.76) | 6.57 (0.36) | 4.32 (1.79) | 2.53 (1.24) | 2.00 (1.41) |
| Probably not | 149 | 4.10 (0.69) | 6.33 (0.78) | 3.60 (1.51) | 2.34 (1.17) | 1.67 (1.15) |
| No | 116 | 3.95 (0.63) | 6.11 (0.95) | 3.65 (1.30) | 2.10 (0.96) | 1.57 (1.08) |
| P value | | **< 0.01** | **0.01** | 0.10 | **0.02** | **< 0.01** |
| Trans coworker | | | | | | |
| Yes | 32 | 4.37 (0.73) | 6.18 (1.07) | 4.05 (1.22) | 3.06 (1.01) | 1.95 (1.40) |
| Probably | 23 | 4.30 (0.86) | 6.34 (0.87) | 4.12 (1.56) | 2.57 (1.45) | 1.89 (1.45) |
| Probably not | 171 | 4.02 (0.72) | 6.23 (0.88) | 3.57 (1.51) | 2.21 (1.11) | 1.74 (1.23) |
| No | 67 | 4.05 (0.54) | 6.36 (0.66) | 3.74 (1.30) | 2.07 (0.99) | 1.52 (1.02) |
| P value | | 0.06 | 0.99 | 0.22 | **< 0.01** | 0.10 |

Note. LGBT-DOCSS-JP total scale scores and subscale scores are described. All statistical tests used the Jonckheere–Terpstra test. Bold font shows significance at $P < 0.05$ in P value line. SD, standard deviation.
